# Development and Evaluation of an AI-Assisted, Privacy-Preserving Surgical Risk Calculator

**DOI:** 10.1101/2025.04.18.25325614

**Authors:** Nathan Wolfrath, Gopika SenthilKumar, Adhitya Ramamurthi, Anai N. Kothari

**Affiliations:** Department of Surgery, Division of Surgical Oncology, Medical College of Wisconsin; Data Science Institute, Medical College of Wisconsin; Bud and Sue Selig Hub for Surgical Data Science, Medical College of Wisconsin

**Author notes:** **Corresponding Author:** Anai N. Kothari, MD MS FSSO, Medical College of Wisconsin Division of Surgical Oncology, 8701 W Watertown Plank Road Milwaukee, Wisconsin 53226.

## Abstract

Large language models (LLMs) have shown capabilities in generating functional code, yet their utility in the development of clinical prediction tools has not been significantly explored. We evaluated GPT-4o’s capability to create a postoperative complication risk calculator similar to the existing National Surgical Quality Improvement Program (NSQIP) risk calculator. This included data preprocessing, predictive modeling, and development of a web application. Synthetic data of a similar structure to the NSQIP dataset was used when communicating with GPT-4o to maintain privacy. 512 lines of Python code were generated across 14 prompts, with one line requiring human editing. The resulting logistic regression models achieved similar Brier scores compared to the original NSQIP risk calculator and demonstrated strong discrimination (C-statistic > 0.75), while slightly underperforming previously reported predictive metrics for some outcomes. Development was completed in three hours. These findings suggest that LLMs can facilitate rapid development of clinical decision support tools, though output still requires human oversight and refinement.

## Introduction

Large language models (LLMs) are an effective tool for generating operational computer code in response to a user prompt [1,2]. To date, the utility of these tools in the development of predictive clinical tools has not been sufficiently evaluated. In this study, we aim to understand the capability of LLMs in this capacity by using an LLM to generate comprehensive code for a postoperative complication risk calculator. This includes data preprocessing, model training, creation of a user-friendly, web-based application, and comparison of performance against a commonly-used risk calculator.

## Methods

OpenAI’s GPT-4o was used for generation of Python3 code. The analytic approach and application were designed based on the American College of Surgeons (ACS) National Surgical Quality Improvement Program (NSQIP) Universal Surgical Risk Calculator [3]. An identical dataset to that used in the original calculator was used as source data, compiled from the NSQIP Participant Use File (PUF) 2009-2012. To preserve data privacy, a synthetic dataset (NSQIP-PUF-TWIN) was created leveraging Copulas [4] for multivariate modeling and sampling of continuous variables, and custom code to simulate categorical variables. All data sent online to GPT-4 was synthetic and included no real patient information, with AI-generated code applied to the real dataset locally. Development included three tasks:

### Data Preprocessing

Encoding of categorical variables, generating columns such as patient BMI and Current Procedural Terminology (CPT) specific risk scores for each complication, and saving the transformed data.

### Model Creation and Evaluation

Training of logistic regression models to predict target complications. Data was split into training (80%) and testing (20%) sets. Input features and performance metrics (C-statistic and Brier Score) were based on those described in the development of the original NSQIP risk calculator. Confidence intervals were estimated using 1000 sample bootstrapping. The models were compared against previously published NSQIP risk calculator performance (prior to recent conversion of the calculator to a machine learning approach).

### Web Application Development

Creation of a web-based interface for entry of patient parameters and viewing calculated probability of each complication.

## Results

1,414,006 patients were included, with 1,131,204 records used for model training. For data preprocessing, 104 lines of code were generated by GPT-4o using four prompts. Four prompts were used to generate 93 lines of code for predictive modeling, and six prompts were used to generate 315 lines of code for the web-based application. Upon review, code was nearly universally correct, with one line (0.19%) requiring human editing. A full transcript of the LLM interactions and generated code are available at https://github.com/AnaiLab/LLMCalculator.

Discrimination of the LLM-generated risk calculator (GPT-NSQIP) was high across all measured outcomes (C-statistic > 0.75). When compared to the published performance of the Universal NSQIP Surgical Risk Calculator, GPT-NSQIP achieved equivalent or superior performance by Brier score across all outcomes except surgical site infection, and similar but inferior performance by C-Statistic for all outcomes except post-operative pneumonia (Table 1).

**Table 1a:** Target variables and model performance metrics for GPT-NSQIP compared to Bilimoria et al. ACS NSQIP calculator. Ranges represent a 95% confidence interval (not reported in original publication)

| **Outcome** | **C-Statistic (GPT-NSQIP)** | **Brier Score (GPT-NSQIP)** | **C-Statistic (NSQIP)** | **Brier Score (NSQIP)** |
| --- | --- | --- | --- | --- |
| Mortality | 0.939 [0.936, 0.943] | 0.0110 [0.0107, 0.0113] | 0.944 | 0.011 |
| Morbidity | 0.809 [0.806, 0.812] | 0.0670 [0.0662, 0.0678] | 0.816 | 0.069 |
| Pneumonia | 0.870 [0.864, 0.875] | 0.0123 [0.0119, 0.0127] | 0.870 | 0.011 |
| Cardiac | 0.876 [0.870, 0.883] | 0.0072 [0.0070, 0.0075] | 0.895 | 0.007 |
| SSI | 0.789 [0.784, 0.794] | 0.0207 [0.0203, 0.0213] | 0.817 | 0.032 |
| UTI | 0.787 [0.781, 0.793] | 0.0161 [0.0157, 0.0166] | 0.806 | 0.014 |
| VTE | 0.793 [0.783, 0.803] | 0.0065 [0.0062, 0.0068] | 0.819 | 0.009 |
| Renal Failure | 0.878 [0.870, 0.885] | 0.0067 [0.0064, 0.0070] | 0.903 | 0.006 |

**Table 1b:** Input variables used for GPT-NSQIP models.

| **Variable** | **Categories** |
| --- | --- |
| Age Group | <65, 65-74, 75-84, >=85 |
| Sex | Male, Female |
| Functional Status | Independent, Partially Dependent, Totally Dependent |
| Emergency Case | Yes, No |
| ASA Class | 1, 2, 3, 4, or 5 |
| Steroid use for Chronic Condition | Yes, No |
| Ascites Within 30 Days | Yes, No |
| Systemic Sepsis Within 48 Hours | None, SIRS, Sepsis, Septic Shock |
| Ventilator Dependent | Yes, No |
| Disseminated Cancer | Yes, No |
| Diabetes | No, Oral, Insulin |
| Hypertension Requiring Medication | Yes, No |
| Previous Cardiac Event | Yes, No |
| Congestive Heart Failure within 30 Days | Yes, No |
| Dyspnea | Yes, No |
| Current Smoker Within 1 Year | Yes, No |
| History of COPD | Yes, No |
| Dialysis | Yes, No |
| Acute Renal Failure | Yes, No |
| BMI | Calculated based on inputted height, weight |
| CPT-Specific Risk | Unique per CPT score and outcome |

Additionally, a functional web-based application was created (Figure 1a/1b). Total development time was 3 hours.

**Figure 1a:**
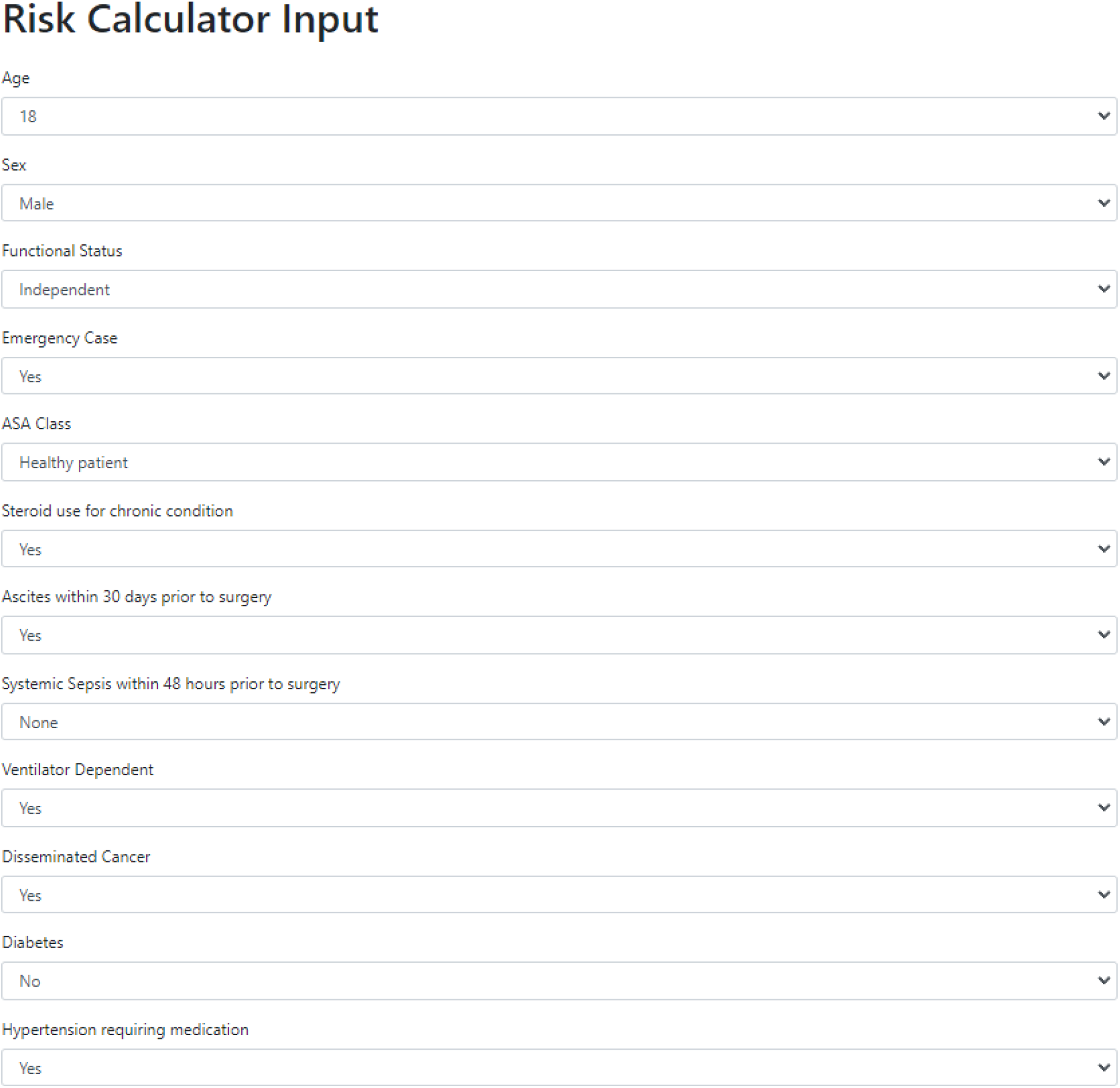
Portion of the input page of the generated application.

**Figure 1b:**
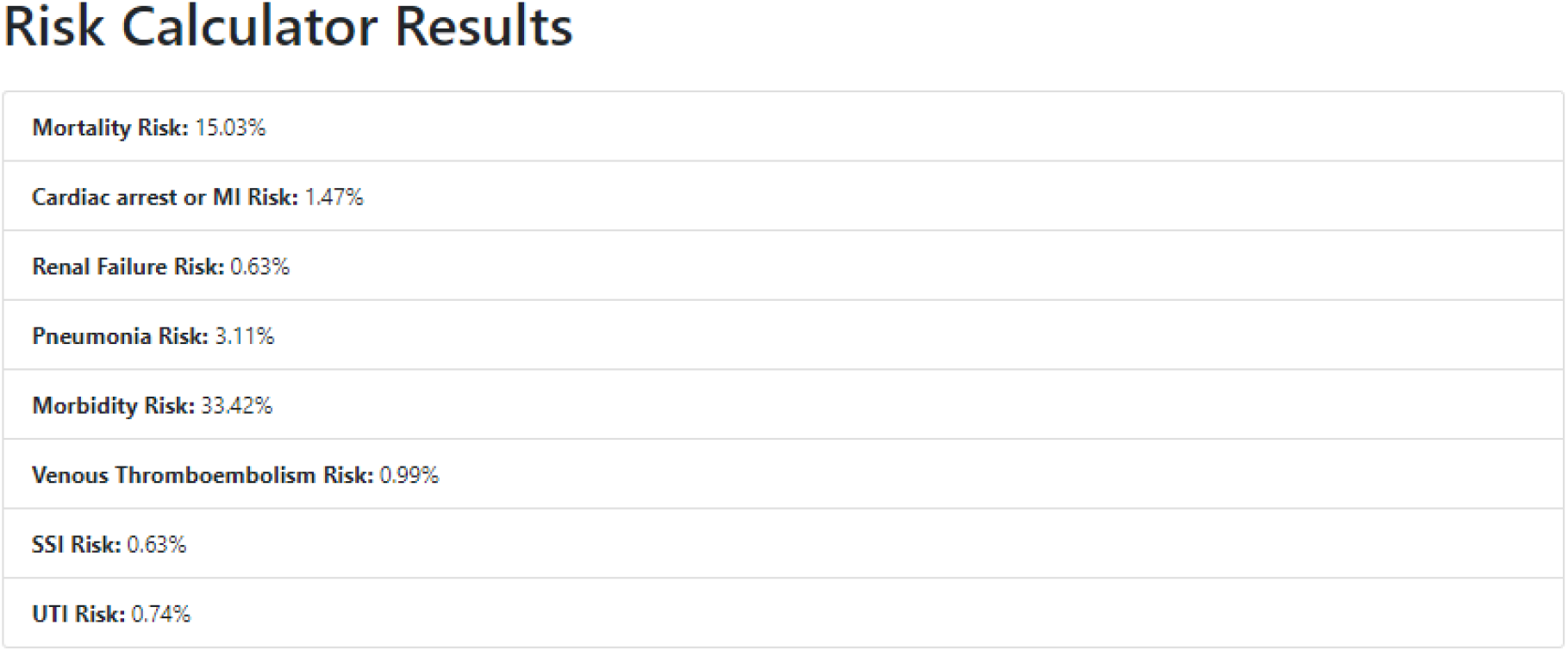
Example results page of the GPT-4 generated application.

## Discussion

Our results indicate that LLMs can assist in the creation of surgical risk estimation tools that achieve similar performance to existing clinically utilized models. All metrics indicate strong predictive performance, and in some cases are superior (i.e. lower Brier score) to previously published results. In areas where underperforming, GPT-NSQIP results still displayed similar results and impressive discrimination. Of note, estimated confidence intervals were not available from the comparison study, which somewhat limits interpretation of metrics. Additional possible reasons for non-identical metrics include differences in training and test dataset splitting, differences in methodology for incorporating procedure-specific risk, and full reliance on LLM-based code with minimal human modification. While the final model and generated application necessitate refinement and human review prior to clinical adoption, the ease and pace of development provides an opportunity for rapid experimentation and development in a variety of healthcare modeling applications.

## Data Availability

Data in this study is from the National Surgical Quality Improvement Program (NSQIP) database and can be requested by investigators at participating institutions at facs.org.

